# Legionnaires’ disease in Switzerland: Rationale and study protocol of a prospective national case-control and molecular source attribution study (*SwissLEGIO*)

**DOI:** 10.1101/2022.10.21.21265377

**Authors:** Fabienne B. Fischer, Melina Bigler, Daniel Mäusezahl, Jan Hattendorf, Adrian Egli, Timothy R. Julian, Franziska Rölli, Valeria Gaia, Monica Wymann, Françoise Fridez, Stefanie Bertschi, the SwissLEGIO Hospital Network

## Abstract

Switzerland has one of the highest annual Legionnaires’ disease (LD) notification rates in Europe (7.8 cases/ 100,000 population in 2021). The main sources of infection and the cause for this high rate remain largely unknown. This hampers the implementation of targeted *Legionella* spp. control efforts. The *SwissLEGIO* national case-control and molecular source attribution study investigates risk factors and infection sources for community-acquired LD in Switzerland. Over the duration of one year, the study is recruiting 205 newly diagnosed LD patients through a network of 20 university and cantonal hospitals. Healthy controls matched for age, sex, and residence at district level are recruited from the general population. Risk factors for LD are assessed in questionnaire-based interviews. Clinical and environmental *Legionella* spp. isolates are compared using whole genome sequencing (WGS). Direct comparison of sero- and sequence types (ST), core genome multilocus sequencing types (cgMLST), and single nucleotide polymorphisms (SNPs) between clinical and environmental isolates are used to investigate infection sources and the prevalence and virulence of different *Legionella* spp. strains detected across Switzerland. The *SwissLEGIO* study innovates in combining case-control and molecular typing approaches for source attribution on a national level outside an outbreak setting. The study provides a unique platform for national Legionellosis and *Legionella* research and is conducted in an inter- and transdisciplinary, co-production approach involving various national governmental and national research stakeholders.

## Background

Legionnaires’ disease (LD) is a severe form of pneumonia with a case fatality of 5-10% [1, 2]. The disease is caused by Gram-negative *Legionella* spp. bacteria, ubiquitously found in freshwater environments and soil. The bacterium is facultative intracellular and replication in amoeba is likely the predominant mechanism for its proliferation. This interaction with amoeba plays an important role in the persistence and release of *Legionella* spp. from its environmental reservoirs [3, 4]. Transmission to humans occurs through inhalation of aerosols or aspiration of water containing *Legionella* spp. In the lung, *Legionella* spp. is phagocytosed into alveolar macrophages, where it replicates intracellularly [5]. Human-to-human transmission is a rare exception [6].

In Switzerland, LD is notifiable to the Federal Office of Public Health (FOPH) [7]. Similar to trends observed in other European countries [8], notification rates for LD in Switzerland continue to rise. In 2021, the notification rates reached a new high of 7.8 cases per 100,000 population [2]. About 70% to 80% of all reported LD cases in Europe, including Switzerland, are community-acquired with the majority of cases occurring sporadically, in contrast to outbreaks or clusters [2, 9]. To date, numerous sources including showerheads, dental units, cooling towers, and fountains have been linked to community-acquired *Legionella* spp. infections (CALD) [10-13], yet little is known about their contribution to the overall disease burden [14, 15].

Estimating the impact of infection sources on the overall disease burden is difficult. In order to draw any significant conclusions on the contribution of a potential infection source to the disease burden, a sizable proportion of LD patients must be screened as LD remains relatively rare.^1^ Regional variability in notification rates additionally suggests that infection sources might differ between regions [17-19], hampering the generalisability of results to different geographic areas. To link a LD case to an infection source, genomic comparison of *Legionella* spp. isolates recovered from patients with LD and from the environment is required. Such molecular epidemiological investigations are resource-intensive and challenging for multiple reasons: First, clinical *Legionella* spp. isolates are recovered from only about 5 to 10% of patients in routine surveillance [2, 8]. Second, the ubiquity of the *Legionella* bacteria and the variable incubation period of 2 to 14 days for LD [5] requires consideration of multiple potential infection sources for a single LD case [20, 21]. The incubation time may also create an inherent delay of up to 14 days between the time a patient is infected and the time environmental samples can be collected. The delay between infection and environmental source investigation might be further prolonged based on reporting timelines set for case notifications by public health authorities.^2^ This prolongation of the period between the time of infection and the investigation of the environmental source may reduce the chances to successfully recover the disease-causing *Legionella* spp. strains from a suspected infection source. Finally, the recovery of *Legionella* spp. isolates may depend on the chosen sampling approaches (e.g. exact sampling location but also procedures) and sampling time points as the detachment of bacteria from biofilms and their release from amoeba may vary over time [22, 23]. Additionally, culture isolation of *Legionella* spp. from environmental samples is labour intensive. It requires careful selection of culture plates and pre-treatment conditions prior to plating (e.g. filter concentration, heat treatment) to optimise growth conditions for *Legionella* spp. and minimise overgrowth of plates by competing organisms. As a result, molecular source attribution of sporadic CALD are primarily reported in single case studies. From such studies is difficult to conclude on a source’s contribution to the overall disease burden [24, 25].

Conducting a combined case-control and molecular source attribution study allows to address some of the challenges outlined above. The case-control study design enables the exploration of various (including transient) host, behavioural, and environmental exposure risk factors for CALD [26-29]. Data obtained from case-control questionnaires can inform the sampling of potential environmental infection sources, in turn, facilitating molecular source attribution [13, 30]. For now, combined case-control and molecular source attribution studies in Europe focused primarily on urban settings or were part of outbreak investigations and, thus, did not investigate any regional variability of infection hazards [13, 30, 31]. The potential of combined molecular and epidemiological approaches to investigate infection sources for sporadic CALD cases at a national and general population level has not yet been realised.

Herein, we present the study design of a national case-control and molecular source attribution study (*SwissLEGIO*). The study aims at investigating risk factors and possible exposure sites for community-acquired, mainly sporadic, LD cases across Switzerland. Engaging with a network of 20 participating university and cantonal hospitals, and collaborating closely with the National Reference Centre for *Legionella* (NRCL), the project creates a framework that enables timely, nationwide recruitment of patients with LD, and facilitates the collection and processing of clinical *Legionella* spp. isolates. Together with the *Legionella Control in Buildings (LeCo)* research consortium [32], environmental source investigations are conducted within days after case detection.

## Methods

### Study design and objectives

This research comprises of a one-year, prospective, national case-control study applying whole genome sequencing (WGS) to link LD patients to potential exposure sources in Switzerland. Host, behavioural and exposure risk factors are investigated by conducting interviews with newly diagnosed LD patients (cases) and healthy control subjects. For patients, further parameters on the clinical, radiological and laboratory characteristics, the clinical case management, disease severity, and health outcomes are extracted from electronic medical records. For a subset of cases and controls, clinical and environmental *Legionella* spp. isolates are collected and sequenced (Fig. 1).

**Fig. 1.**
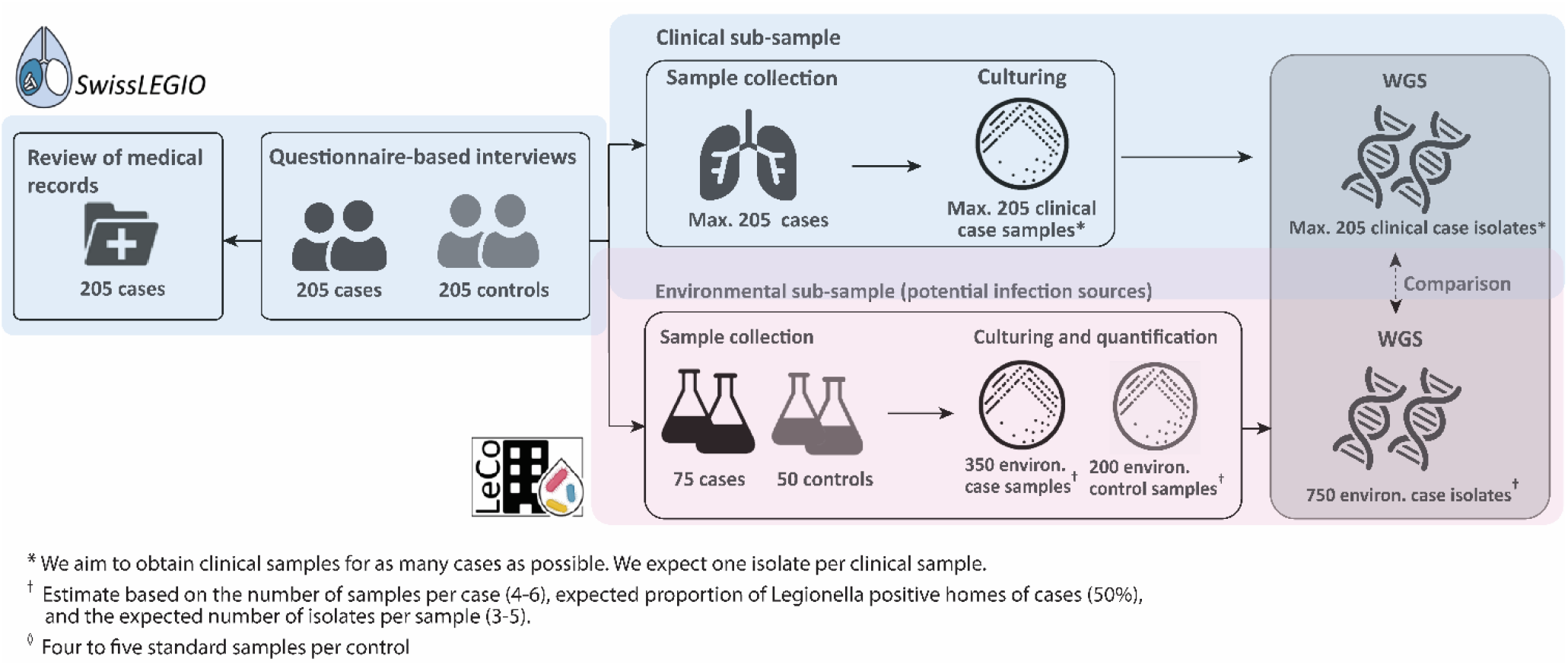
Study design for the national case-control and molecular source attribution study on Legionnaires’ disease in Switzerland, *SwissLEGIO*. The environmental sampling and sample analytics is conducted in collaboration with the *LeCo* Consortium

The objectives of the *SwissLEGIO* study are: (i) To identify host, behavioural, and environmental risk factors for LD, (ii) To attribute infection sources to LD cases by comparing clinical and environmental *Legionella* spp. isolates using WGS, (iii) To assess the genome sequence of *Legionella* spp. differing in virulence and to identify potential traits of more virulent strains, (iv) To assess strain diversity and concentration of *Legionella* spp. in standard household and other environmental samples,^3^ (v) To explore the illness experiences of patients with LD, their health-seeking and long-term quality of life, and (vi) To describe clinical, laboratory, and radiological characteristics of LD and the patient’s clinical case management.

The study involves multiple governmental and research stakeholders. Since its inception, the FOPH, the NRCL, and the Federal Food Safety and Veterinary Office (FSVO) are involved as advisory and strategic planning partners. For the implementation, we closely collaborate with a hospital network consisting of 20 university and cantonal hospitals, the NRCL, the Institute of Medical Microbiology (IMM) at the University of Zurich, and the *LeCo* consortium led by Eawag^4^ [32].

### Study setting, recruitment process and participation eligibility

#### Cases of Legionnaires’ disease

The study includes newly diagnosed LD patients from all of Switzerland over a one-year period to account for seasonal and meteorological impacts on infections [33, 34]. Patients are recruited through a hospital network representing a significant proportion of diagnosed LD patients (the network collectively reported about 55% of all LD cases between 2018 and 2020) (Fig. 2). The decision to recruit through hospitals and the selection of the participating hospital sites were informed by previous research: in Switzerland, diagnostic testing for Legionella spp. is mainly limited to the hospital setting and, therefore, most reported cases are identified at the hospital. In outpatient care, patients with pneumonia are primarily treated empirically or are referred to the hospital for further clinical and diagnostic evaluations [35]. An in-depth analysis of LD notification data in Switzerland was used to identify hospitals notifying the most LD cases and also showed that LD notification rates regionally differ across Switzerland [2], highlighting the importance to include patients from all seven greater regions (NUT-2 level).

**Fig. 2.**
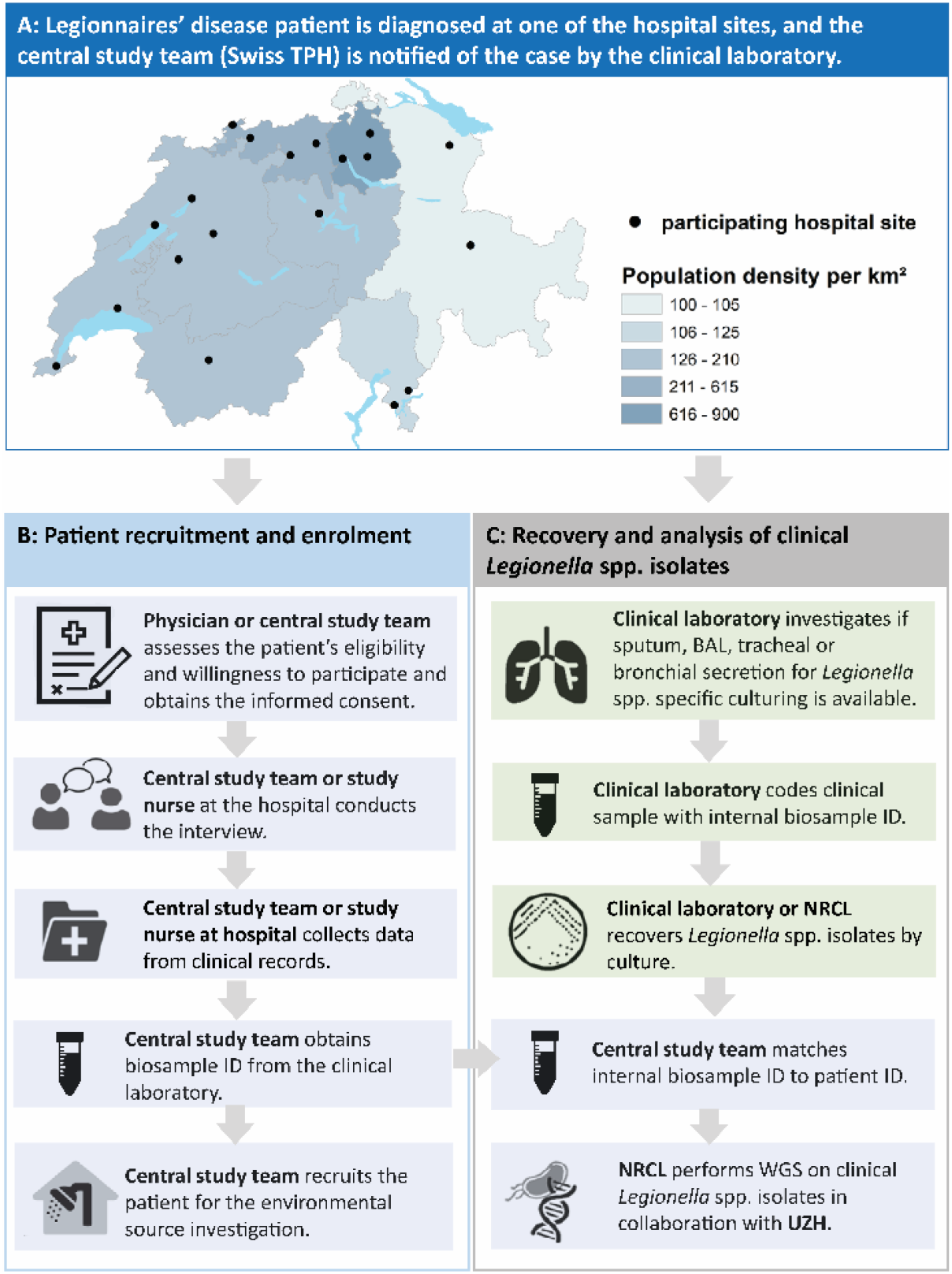
*SwissLEGIO* operational flowchart for the recruitment and the collection of data from patients with Legionnaires’ disease: a) Overview of the *SwissLEGIO* hospital network, b) Data collection upon enrolment, c) Collection and analytics for clinical samples as part of routine case management. BAL: Bronchoalveolar lavage; NRCL: National Reference Centre for *Legionella*. WGS: Whole Genome Sequencing; UZH: University Hospital Zurich

For participating hospitals, individualised recruitment procedures were developed to ensure that the study is embedded optimally in each hospital’s existing workflows and to help minimise the risk of missing any admitted patients with LD. In brief, the central study team at the Swiss Tropical and Public Health Institute (Swiss TPH) is immediately informed by the hospital’s clinical laboratory in case of a positive diagnostic *Legionella* spp. test result. The central study team then coordinates with the hospital’s appointed study physician or the attending physician on the pre-assessment of the patient’s eligibility for participation (the eligibility criteria are summarised in Table 1) and the subsequent enrolment. Written study-specific informed consent is obtained before the questionnaire-based interview is conducted by the study physician, the study nurse, or the central study team (Fig. 2).

**Table 1.**
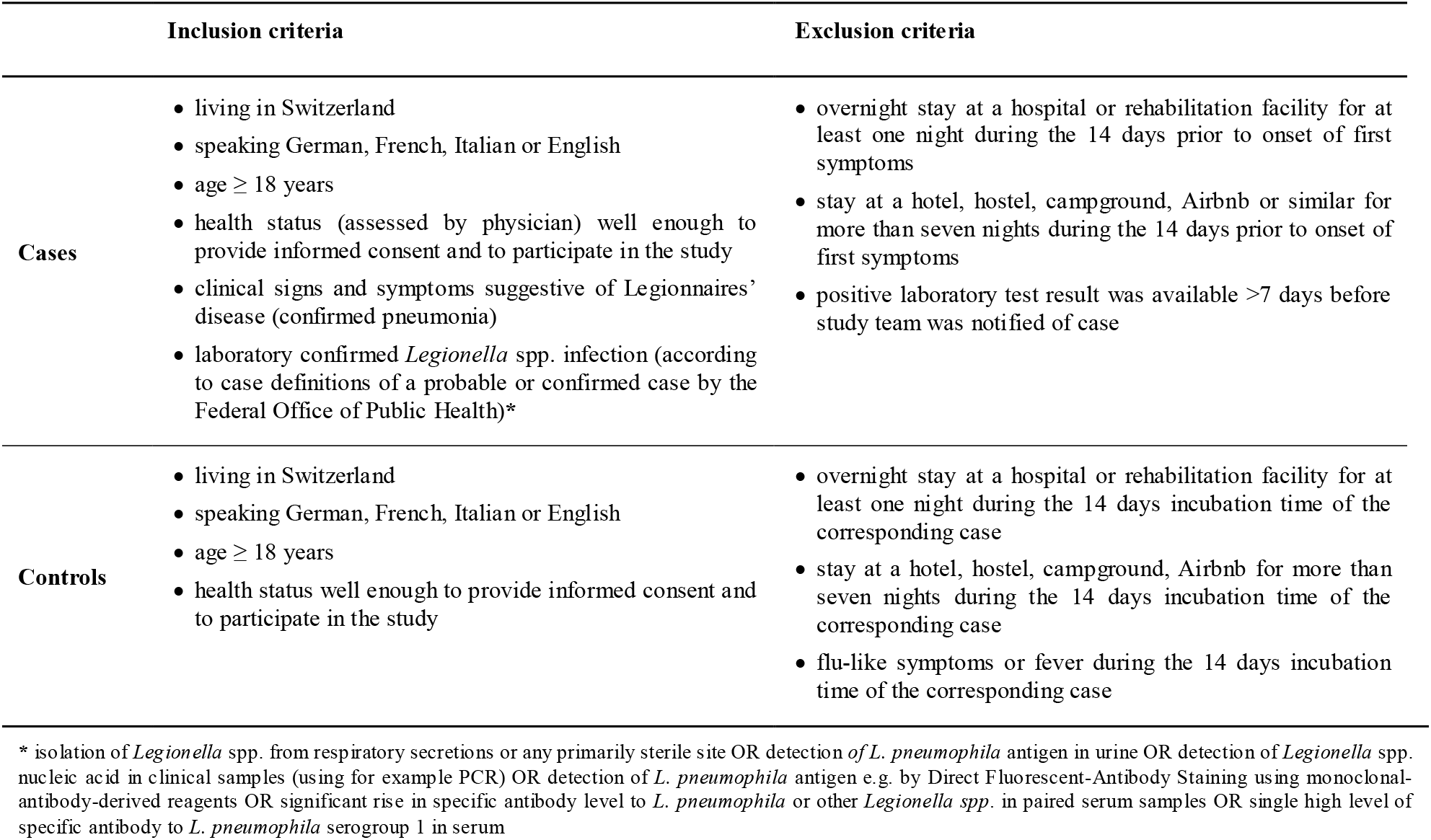
Summary of eligibility criteria (inclusion and exclusion) for participation in *SwissLEGIO*

### Controls

One control per enrolled case is recruited from the general Swiss population. Controls are selected from a dataset based on the national census list, which comprises a random population sample. The dataset is provided by the Swiss Federal Statistical Office [36]. A control matched to a LD case for age (+/-5 years), sex, and location of residence (district level/ “Bezirksebene”) is chosen and contacted by e-mail or postal mail as soon as a case has been enrolled. Following the written invitation, the study team assesses the control’s eligibility (Table 1), ability and willingness to participate in the study by phone. Informed consent from controls is obtained prior to the interview. Our research group has successfully applied this recruitment approach in a previous study [37].

### Sample size calculation

Calculation of the sample size was performed using Epi Info™ 7 (Centers for Disease Control and Prevention, USA). We consider the ubiquitous nature of *Legionella* spp. in the environment and assume that 60% of controls are exposed to a risk factor during the period of potential risk exposure [30, 38]. Therefore, a sample size of 205 cases and 205 controls is required to detect an odds ratio (OR) of 2 with 90% power and alpha=0.05. We adjusted the size of the hospital network to reach the required sample size within one year.

### Data collection and piloting

For cases, data and biological samples are collected at three different time points: (i) the treating physician obtains clinical samples suitable for *Legionella-*specific culturing prior to enrolment of the patient in the study, (ii) after informed consent is obtained, a questionnaire-based interview on potential risk exposures is conducted and electronic medical records are reviewed, (iii) thereupon environmental samples from potential risk exposure sites are collected for a subset of cases. Preference for the environmental sampling is given to cases from whom clinical *Legionella* spp. isolates are available. For controls, the questionnaires-based interview is conducted and environmental samples are collected from a subset of controls matched to a LD case (for whom environmental samples were also collected) (Fig. 1).

Data and biological sample collection were carefully piloted in a two-step approach: in a first step, between October 2020 and October 2021, direct recruitment of patients with LD through the hospital was tested in collaboration with the University Hospital Basel. Moreover, by interviewing newly diagnosed LD patients, the manageability and comprehensibility of the questionnaire-based case-control interview were assessed. In a second piloting step, from March to June 2022, the participant invitation process, the electronic data collection tools, the coordination of the case-control interview, and the subsequent environmental sample collection, shipment, and analysis were pretested with healthy volunteers. Additionally, data collectors and laboratory staff of the central study team were trained on the data collection and laboratory processing of environmental samples. Finally, the manageability and comprehensibility of the interview completion guidelines and study-specific standard operating procedures (SOPs) on participant recruitment, sampling and sample analysis were tested.

### Clinical samples

Clinical samples suitable for *Legionella*-specific culturing (such as sputum, Bronchoalveolar lavage (BAL), tracheal or bronchial secretion, or pleural fluid in case of pleural effusion) are collected by the treating hospital physician as part of the routine clinical management of the patient and, hence, prior to patient enrolment. The hospitals’ clinical laboratories isolate *Legionella* spp. by culture on charcoal-based agar and subsequently send the isolates to the NRCL as described in the guidelines for notifiable infectious diseases from the FOPH [7]. If a clinical laboratory cannot perform *Legionella-*specific culturing, clinical samples can be sent directly to the NRCL for further processing (i.e. culturing and serotyping). Upon enrolment, the study team enquires if *Legionella* spp. could be isolated from clinical samples and obtains written informed consent from the patient for the use of the *Legionella* spp. isolates in the study.

*Legionella* spp. isolates are sent from the NRCL to the IMM for WGS analysis (Fig. 2). We expect to analyse one *Legionella* spp. isolate per patient [39].

### Case-control questionnaire and patient records

Upon completion of the informed consent, the central study team or a study nurse conducts questionnaire-based interviews with LD patients and controls. The questionnaire is based on information from published LD data collection tools (either for routine assessment [40, 41] or other case-control studies [27, 30]) and current literature on risk factors and exposure sites for LD. Swiss federal stakeholders in *Legionella* spp. control namely the FOPH, the FSVO, the Federal Office of Energy (SFOE) and collaborating researchers from the *LeCo* consortium were consulted for inputs on the questionnaire design.

The questionnaire consists of 20 sections and focuses on the 14 days before the onset of illness for cases and the same (matched) time period for controls. The questionnaire covers potential predisposing host risk factors for LD (e.g. age, sex, co-morbidities), potential behavioural risk factors for LD (e.g. regularly showering at sports facilities, gardening habits) and investigates exposure to potential environmental infection sources (e.g. housing water installation, public artificial water sources, natural water sources). For cases, the questionnaire further covers the illness experiences of patients with LD and their health-seeking (Table 2). Pretested interviews lasted approximately 60 minutes and were well received by patients and healthy control volunteers in content, length, and flow.

**Table 2.**
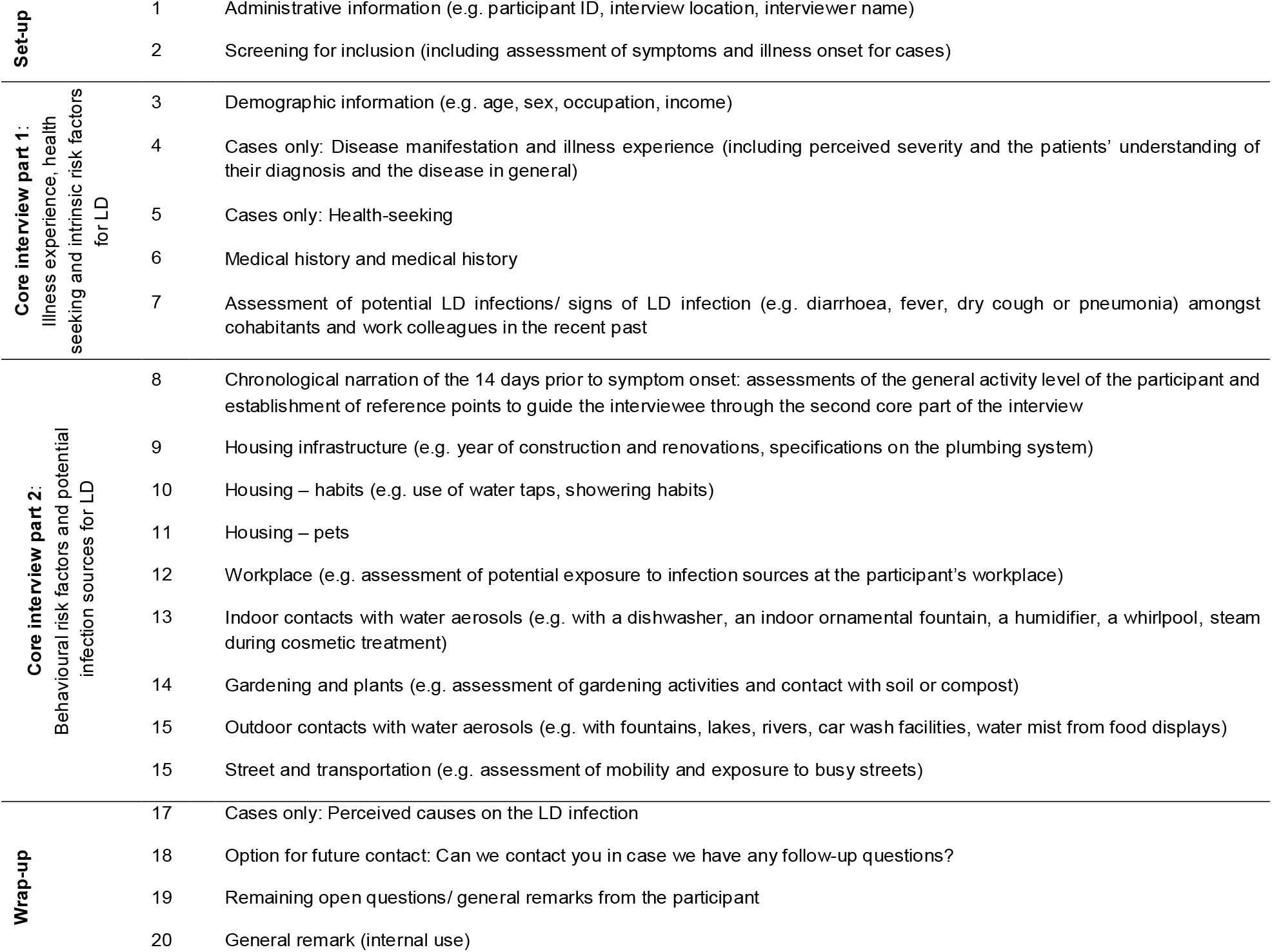
Structure of the *SwissLEGIO* case-control questionnaire for Legionnaires’ disease

For cases, parameters on the patients’ clinical case management and the disease severity are extracted from the electronic medical records. The parameters include the medical history, the timespan between onset of symptoms and admission to the hospital, length of hospital stay, CURB-65 parameters [42], radiological findings (e.g. consolidation, crazy paving, bronchial wall thickening, pleural effusions), ICU admission and length of ICU stay, disease progression within 48h after admission, performed diagnostics, laboratory parameters and prescribed treatment.

### Environmental samples

Environmental samples and information on residential buildings and water installation attributes for a subset of participants are collected by the central study team. Up to six environmental samples from 75 cases within 14 days following the questionnaire-based interview are collected (Fig. 1). Up to five of these samples are standardised water samples collected in the patient’s home: (i) first flush (1 litre, mix of cold and warm water) kitchen tap water (ii) first flush (1 litre, mix of cold and warm water) shower head water from the most used shower, (iii) sequential sample from the hot water line collected at the most-used shower/ bathtub^5^, (iv) sequential sample from the cold water line collected at the shower/ bathtub^6^ of the most-used shower, and (v) first flush (1 litre, mix of cold and warm water) from the second-used shower. The detailed procedures for water sampling are found in the supplementary file). Prior to this sampling, the participant is instructed to refrain from using the taps for four hours. In addition to the standard samples, up to two samples are collected from other likely environmental risk exposure sites reported in the patient interview. These exposure sites are sampled from private locations (e.g. garden hose, water dispenser or humidifier) or public locations (e.g. spas, car wash facilities, decorative fountains, air-conditioners or cooling towers on hotels or supermarkets in proximity of patient’s residency, permissions provided). For a sub-set of 50 healthy controls, only standard household water samples are collected (Fig. 1).

Environmental samples are cultured and the number of colony forming units (cfu) of *Legionella* spp. in the original water sample are estimated according to ISO 11731 guidelines. For the ISO culturing of standard household water samples, we prepare three plates per sample using the following pre-treatment conditions: filtration only, filtration plus heat treatment, and filtration plus acid treatment. Culture plates are regularly checked for growth for one week and up to three suspected *Legionella* spp. colonies of each morphology are culture-confirmed by direct plating of the colony on charcoal agar plates with and without L-cysteine. Isolates showing no growth on plates without L-cysteine are considered *Legionell*a spp. (Fig. 3a). All culture-confirmed *Legionella* spp. isolates will be characterised by MALDI-TOF mass spectrometry (MS) (to differentiate strains) and agglutination tests (to differentiate serogroups of *L. pneumophila*). All *Legionella* spp. isolates recovered from potential infection sources of cases that are matching the strain and/ or serogroup of the case’s clinical *Legionella* spp. isolate are sent to the IMM for further strain characterisation using WGS (Fig. 3b). Finally, flow cytometry (for total cell count), IDEXX Legiolert (quantification of culturable *L. pneumophila)*, and digital PCR (quantification of the *ssrA* gene for *Legionella* spp., *mip* gene for *L. pneumophila* and the *wzm* gene specific to *L. pneumophila* serogroup 1) are performed for quality control (Fig. 3c). The digital PCR protocol was developed by adapting the qPCR assay from Benitez and Winchell [43].

**Fig. 3.**
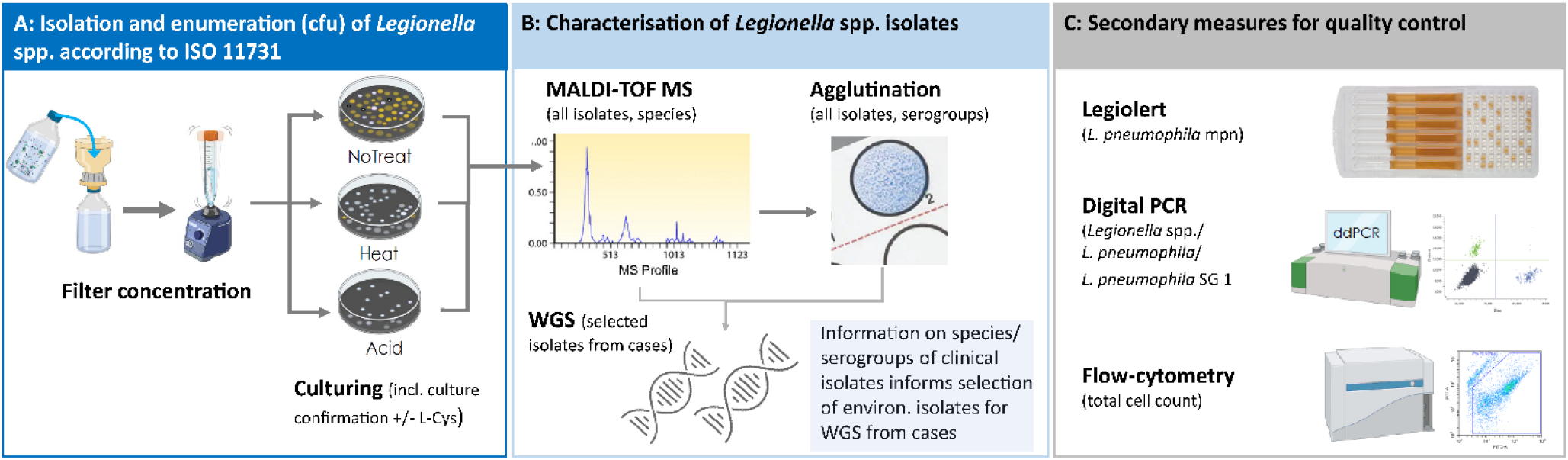
Overview of the laboratory analytics pipeline for the isolation and characterisation of environmental *Legionella* spp. strains from standard household and other environmental samples: a) Summarises the isolation and enumeration of *Legionell*a spp. according to ISO 11731, b) Culture-confirmed *Legionella* spp. isolates are characterised by MALDI-TOF MS and agglutination tests and are selected for WGS, c) For quality control, flow-cytometry, digital PCR and Legiolert are performed for all samples. cfu: colony-forming units; L-Cys: L-Cysteine; MS: mass spectrometry; WGS: Whole Genome Sequencing; SG: serogroup; mpn: most probable number

For additional samples from potential exposure sources of patients, sample processing approaches are assessed on a case-by-case basis in consultation with researchers and external research partners from the *LeCo* consortium. The characterisation pipeline for such isolates will remain the same as for the standard household water samples. Based on rough estimates from the literature [12, 30, 44], we expect to sequence a total of 750 environmental *Legionella* spp. isolates recovered from potential exposure sources of cases (Fig. 1).

### Data management

Data is collected on standardised electronic Case Report Forms (eCRF) using the data collection software Open Data Kit (ODK, getodk.org). Forms are identified by subject IDs. Automated validation tools in the eCRF check for data completeness and plausibility during data entry. During data collection, the data collected is continuously checked by the research team for completeness, plausibility, and accuracy. Additionally, random source data verification is performed. Data is stored on a secured network drive accessible only to authorised study team members. Data on the network drive is backed up regularly, according to Swiss TPH institutional policy. Radiological images are coded and securely shared between hospitals via a secure data exchange platform, and the images are securely stored on two password protected hard drives.

Quality control measures for the analysis and storage of biological samples are performed according to the study laboratories’ routine standard operation procedures.

## Statistical methods and analysis

### Epidemiological analysis

Cases and controls are characterised in terms of demographics, illness experience (only cases), health-seeking (only cases), and comorbidities. Crude OR for LD will be calculated by running univariable logistic regressions on single risk factors. Based on results of the univariable logistic regression and biological or epidemiological plausibility, variables will be subsequently selected for a multivariable (unconditional) regression to calculate adjusted OR (aOR). The population attributable fraction (PAF) is calculated for each statistically significant risk factor of the multivariable model as the difference of observed cases and expected cases in absence of the risk factor. The analysis will be conducted with the statistical software R [45]. Potential exposure sites of cases will be geocoded using geographic information systems (GIS) to assess regional distributions of LD cases and to identify clustering of potential infection sources.

Additionally, analysis of radiological imaging is performed independently by at least two experienced radiologists. All chest X-rays or CT scans are evaluated blinded from clinical and microbiological information for lung involvement, distribution (e.g. upper vs. lower lobes, uni- versus multilobar), radiological patterns (e.g., consolidation, ground glass opacities, cavitation, nodules, crazy paving, bronchial wall thickening), pleural effusion, and lymphadenopathy.

### Analysis of biological samples

WGS is performed using the IMM’s internal ISO accredited (ISO/IEC 17025) sequencing workflow and analytical pipeline for the characterisation of the *Legionella* spp. strains. This workflow and the analytical pipeline are already in use for the characterisations of *Legionella* spp. isolates currently performed for the NRCL [7]. For library preparation, the Illumina NextFlex assay is used and sequencing is performed batch wise at a NextSeq 1000i with 150nt paired end sequencing. After sequencing, the data is quality controlled, raw reads are assembled, and analysed using SeqSphere (Ridom) and the CLC workbench. Only genomes with an average minimal coverage of 40-fold or more will be further evaluated. The sequence type (ST) of all isolates will be determined and the genomic relatedness will be visualised with a series of phylogenetic tools such as core genome multi-locus sequence typing (cgMLST) e.g. as neighbour joining tree or as a single nucleotide polymorphism (SNP) tree. Fig. 4 illustrates such visualisation for a *Legionella* spp. outbreak investigation conducted in Basel: allelic differences between *L. pneumophila* isolates shown in the figure are based on a published cgMLST scheme for *L. pneumophila* using 1’521 allelic loci [46].

**Fig. 4.**
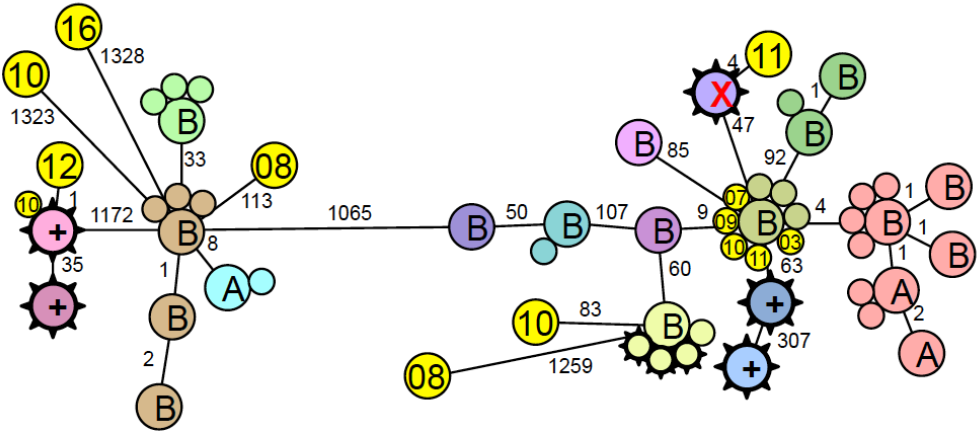
Environmental source of *L. pneumophila*. Isolates labelled with A and B are from two air conditioning cooling towers. Isolates with spikes are human isolates from the same seasons. Circles with numbers are human isolates from previous years. The small numbers between the circles indicate the number of allelic differences between two isolates (data from egli-lab, IMM)

All successfully sequenced strains will be contextualised with previously sequenced isolates from the Swiss database and with global available sequences from public data repositories such as the National Center for Biotechnology Information (NCBI). In addition, we will compare the human and environmental isolates using a bacterial genome wide association study approach. We will link clinical phenotypes (e.g. LD, disease severity) to potential enriched sequence types, genes, k-meres, or SNP in isolates causing invasive disease [47]. We aim to identify genes and annotate potential functionalities linked to the clinical phenotypes. All sequenced genomes are shared according to FAIR principles [48] on the Swiss Pathogen Surveillance Platform (www.spsps.ch). The usage of the platform further eases data exchange between the different research partners [49].

## Strengths and limitations

The *SwissLEGIO* study enrolls patients with LD from all seven greater regions of Switzerland. To our knowledge, this is the first national case-control and molecular source attribution study that is conducted outside of an outbreak setting in Switzerland. The study design innovates in (i) minimising the timespan between LD symptom onset and enrolment of cases in the study, (ii) enhancing and promoting the collection of clinical *Legionella* spp. isolates, and ensuring a high sensitivity of the molecular source attribution approach. All three aspects are crucial to ensure a successful linking of clinical isolates to an infection source. The direct recruitment of cases through an established hospital network instead of the National Notification System -as in previous studies [26, 27] -significantly reduces the timespan between the patient’s diagnosis and enrolment in the study. When piloting the recruiting process, the time between patient’s hospitalisation and case notification to the study team averaged at three days. This is significantly shorter than the Swiss legal requirements to report an LD case within seven days to the National Notification System [7]. The recovery of clinical *Legionella* spp. isolates was a major limitation in environmental source investigations in previous studies [20, 50]. By directly recruiting LD patients through the hospital network, and by exchanging closely with hospital partners on an ongoing basis, we believe to address this challenge by promoting and facilitating the collection of clinical *Legionella* spp. isolates.

We ensure a high sensitivity of the molecular source attribution approach by applying WGS, which has a strong discriminatory power between different *Legionella* spp. strains and, therefore, allows a direct comparison of environmental and clinical isolates [50]. During LD outbreak investigations, WGS analysis was successfully used to trace clinical *Legionella* spp. strains in the environment [12, 51, 52]. Yet, such outbreak investigations also highlighted the complexity of developing sensitive environmental sampling, culturing and isolate selection strategies to account for high *Legionella* spp. strain diversities in environmental samples [12, 31]. In addition, several outbreak studies do not detail on their methods applied for the recovery and selection of environmental *Legionella* spp. isolates [31, 51, 52]. Therefore, the challenges of obtaining and subsequently selecting appropriate environmental *Legionella* spp. isolates for WGS in a streamlined manner and the applicability of WGS for the investigations of sporadic LD cases remain largely unexplored. For the *SwissLEGIO* study we developed streamlined processes for the collection and processing of standard environmental household and other environmental samples and carefully implemented *Legionella* spp. isolates characterisation measures that will inform the selection of isolates for WGS analysis. Finally, the *SwissLEGIO* study supplements the sequencing data with a rich set of epidemiological metadata. This epidemiological context information is essential for the interpretation of observed similarities between different environmental and clinical *Legionella* spp. strains and hence for infection source attribution [50, 53].

Data collected during the questionnaire-based interviews might be subject to reporting biases. The selective memory effect of participants (recall bias) was accounted for by the design of the questionnaire: during the interview, the participant is guided in a structured manner through a wide range of potential exposure sources. To improve recall, participants are interviewed as soon as possible after the LD diagnosis is confirmed. By using an established hospital network, the time between diagnosis, interview and environmental sampling of risk exposures can be kept short. Additionally, any cases for which the diagnostic assessment occurred more than seven days before the study team is notified are excluded. The number of environmental samples collected and isolates analysed is limited due to financial constraints. Some non-standardised environmental samples may not be collected if access is difficult and/or permissions cannot be obtained.

## Future perspectives and impact on policy

Large-scale combined molecular and epidemiological studies are sorely needed to identify infection sources of CALD and to explore their risk magnitude and regional variability. Such studies are essential to understand the observed increase in LD notification rates and to improve prevention efforts in general and in Switzerland in particular. The *SwissLEGIO* study aims at addressing these needs. The study draws on a unique range of scientific and policy expertise on *Legionella* spp. and LD in Switzerland. Through close collaboration with national partners in aquatic science and building-technology research, we harness expertise in environmental sampling and sample analysis, bridging the gap between human health and environmental exposure. The recruitment through the hospital network and the close collaboration with the NRCL and IMM aligns the study with *Legionella* spp. surveillance activities enacted by the FOPH [7]. The support from and regular consultations with the Federal Offices (FOPH and FSVO) further ensures the project aligns with governmental research needs, facilitating research uptake and policy improvement.

The study contributes to capacity building for future national *Legionella* surveillance and LD case management. Through the hospital network, the study raises awareness for LD and promotes the collection and analysis of clinical *Legionella* spp. isolates as part of the routine surveillance. Clinical, radiological and laboratory data from the *SwissLEGIO* study will be used in satellite projects to validate the *Legionella* score developed by Fiumefreddo *et al*. [54] and to systematically assess radiological characteristics of pneumonia caused by *Legionella* spp. in a large LD patient population that is representing a significant proportion of reported LD cases in Switzerland. For both of these assessments we will use a control group of suspected pneumonia cases tested negative with the *Legionella* Urinary Antigen Test. This analysis of clinical, laboratory and radiological characteristics of LD together with data that is collected on patients’ health seeking and recovery from LD may inform revisions of current pneumonia management guidelines. Experiences gained from processes established during this study will also aid the effort to introduce a nationally standardised questionnaire for future case and outbreak investigations. As of today, Switzerland lacks such a comprehensive LD outbreak investigation toolbox [15]. This results in procedures applied to address LD clusters or outbreaks within Switzerland being heterogeneous and, hence, the responsibilities of different stakeholders not being well defined [12, 15, 55]. In turn, this hampers a successful and timely detection of the cluster’s infection source. Lastly, the study will, as part of the *LeCo* consortium’s research portfolio, play an important role in assessing and informing stakeholders and authorities of the applicability of WGS for single case and cluster investigations during routine surveillance activities.

The *SwissLEGIO* study also provides a unique platform for future research on *Legionella* and LD, including a more in-depth exploration of the bacteria’s complex ecology, of virulence factors, of clinical and laboratory characteristics and also on disease progression and long-term sequelae of LD. Additionally, the *SwissLEGIO* study data is contributing towards the establishment of a nationally centralised biobank for clinical and environmental *Legionella* spp. strains and associated epidemiological metadata on the spsp.ch platform. Similar to EpiPulse, which is currently implemented by the European Centre for Disease Prevention and Control (ECDC) [56], the spsp.ch platform will allow researchers and policy makers to exchange epidemiological and genomic data on LD. Such platforms promote research on LD to be conducted in inter-and transdisciplinary collaborations that are highly needed to address the complex pathway from environmental exposure to *Legionella* spp. to the clinical presentation of LD. Finally, the experiences gained conducting this study, and the data foundation *SwissLEGIO* is providing on LD may provide an opportunity to link Switzerland through LD on a scientific level to international data sharing initiatives and EpiPulse, which connect the European research and public health community.

## Supporting information

Protocol for sampling of standard household water samples

## Data Availability

Not applicable, as the manuscript constitutes a research protocol.

## Acknowledgments

We thank William Rhoads and Frederik Hammes for their valuable advice during the development and validation of the sampling and analytics protocols. We also would like to acknowledge the valuable inputs from all other *LeCo* consortium members supporting the implementation of these protocols. We also thank the cantonal laboratory Basel-Stadt for sharing their experiences on sampling and sample analytics approaches applied by the lab for routine surveillance and source investigations. At the Federal Office of Public Health, we gratefully acknowledge the various inputs and discussions surrounding the case-control study with Sabine Basler, Marianne Jost, Simone Graf, Nicole Gysin, Mirjam Mäusezahl-Feuz, Ornella Luminati and Ekkehardt Altpeter. We also thank the University Hospital Basel (USB) namely Sarah Dräger and Michael Osthoff for supporting the piloting study that informed the *SwissLEGIO* study. We acknowledge the Federal Statistical Office for providing the data for the recruitment of controls from the general population.

## Statements and Declarations

### Ethics approval

Ethical approval for the study was obtained from the Ethics Commission of Northwestern and Central Switzerland (EKNZ, 2022-00880). This study is conducted in accordance with the principles of Good Epidemiological Practice [57] and the Declaration of Helsinki [58]. Data is stored in concordance with Swiss data protection laws.

## Footnotes

1 Despite the strong increase in notification rates, only 678 cases were reported in Switzerland in 2021 [16]

2 Currently 7 days in Switzerland [7].

3 A primary objective of the *LeCo* consortium

4 Eawag – Swiss Federal Institute of Aquatic Science and Technology, ETH Zurich

5 10 times 100ml, flow-proportional: 1 litre representative sample from 10 litre hot water line after flushing for 5s prior to taking first sequential sample

6 10 times 100ml, flow-proportional: 1 litre representative sample from 10 litre cold water line after flushing for 5s prior to taking first sequential sample

