## Supplementary material for "Legionnaires’ disease in Switzerland: Rationale and study protocol of a prospective national case-control and molecular source attribution study (*SwissLEGIO*)": Protocol for sampling of standard household water samples

<u>Prior to sampling</u>

The participant is asked to refrain from using the taps in the kitchen and bathroom for 4 hours prior to the collection of samples.

<u>Kitchen tap; collection of first liter; cold and warm water mixed</u>

- ❒ Determine how cold and hot water are mixed
- ❒ With an open 1-L glass bottle with narrow opening directly below the outlet, open the cold and hot outlet(s) to get approximately equal proportion of cold and hot water flowing through the outlet and fill the bottle

  *Note: Open the outlet to medium flow rate (it should take approx. 5 s to fill the sample bottle)*
- ❒ Remove the sample bottle from the flow (set aside) and immediately collect 250 mL in the 1000 mL plastic beaker (you may quickly turn off the water in between)
- ❒ Turn off the fixture
- ❒ Firmly close the sampling bottle, turn bottle twice upside down to mix the thiosulfate
- ❒ Read and record sample temperature in the beaker with 250 mL (highest temperature that is reached on the display)

<u>Most used shower; collection of first liter from (most used) shower **through the showerhead**; cold and warm water mixed</u>

- ❒ Determine how cold and hot water are mixed
- ❒ <u>Do not</u> remove the shower head from the shower hose
- ❒ If the shower head is too large to direct all the water into the 1-L wide-mouth glass bottle, use a UV irradiated plastic bag to act as a funnel to direct the water into the sampling bottle
- ❒ With the shower head (or funnel if needed) directed into an open 1-L wide-mouth glass bottle, open the cold and hot outlet(s) to get approximately equal proportion of cold and hot water flowing through the outlet and fill the bottle

  *Note: Open the outlet to medium flow rate (it should take approx. 5 s to fill the sample bottle)*
- ❒ Switch the shower head from the 1 L sample bottle to the 1000 mL beaker and collect 250 mL (you may quickly turn off the water in between)
- ❒ Turn off the fixture
- ❒ Firmly close the sampling bottle, turn bottle twice upside down to mix the thiosulfate
- ❒ Read and record sample temperature in the beaker with 250 mL (highest temperature that is reached on the display)

Most used; composite sample of 100 ml from every subsequent 1L from the cold water line **through the existing shower hose**

- ❐ Remove the shower head
- ❐ Set water handle to cold position
- ❐ Turn on the tap and flush for 5 seconds, adjust flow rate so that a 100 mL bottle can be filled without splashing.
- ❐ Take sequential 100 mL samples – repeat 10 times in total so that a composite of 1 L water is collected in one autoclaved 1-L wide-mouth glass bottle:
  - a) Collect 100 ml cold water via shower hose into one of the 100 mL autoclaved glass bottles.
  - b) Move the hose to the plastic beaker and immediately collect 900 mL into the beaker
  - c) Turn off the cold water
  - d) Transfer the 100 mL from the 100 mL bottle to the autoclaved 1-L wide-mouth glass bottle
  - e) **For the 1st, 2nd and 4th liter**: Measure and record water temperature in the beaker
- ❐ Firmly close the sampling bottle, turn bottle twice upside down to mix the thiosulfate
- ❐ Turn on the cold water to the maximum flow rate. Continue flushing cold water until water temperature no longer changes.

  *Note: Coldwater temperature may continually decreases for long periods. Therefore flush the cold water until it is not changing by more than 0.1 °C for approx. 20 s or after a total time of 2 minutes flushing (whichever occurs first) and document it*
- ❐ measure and record the amount of time required to reach steady temperature
- ❐ measure and record water temperature at steady state
- ❐ Continuing flushing the cold water (meaning: do not turn off the water between temperature and flow rate measurements) and measure the water flow rate: record the amount of time it takes to fill the 1000 mL (repeat to have 3 measurements in total)

Most used shower; composite sample of 100 ml from every subsequent 1 L sample from the hot water line

- ➔ If the shower hose was collected, this sample will be collected through the spout where the existing shower hose was installed.
- ➔ If the shower hose was not collected, this sample will be collected through the existing shower hose

- ❐ Set water handle to hot position
- ❐ Turn on the tap and flush for 5 seconds, adjust flow rate so that a 100 mL bottle can be filled without splashing.
- ❐ Take sequential 100 mL samples – repeat 10 times in total so that a composite of 1 L water is collected in one autoclaved 1-L wide-mouth glass bottle:
  - f) Collect 100 ml hot water into one of the 100 mL autoclaved glass bottles.
  - g) Move the hose to the plastic beaker and immediately collect 900 mL into the beaker

    *Note: Here you can increase the flowrate when filling the beaker with 900 mL water to save some time during the sampling,*
  - h) Turn off the hot water
  - i) Transfer the 100 mL from the 100 mL bottle to the autoclaved 1-L wide-mouth glass bottle
  - j) **For the 1st, 2nd, 4th and 10th liter:** Measure and record water temperature in the beaker

- ❒ Turn on the hot water to the maximum flow rate. Continue flushing hot water until water temperature no longer changes.

  *Note: Flush the hot water until it is not changing by more than 0.1 °C for approx. 20 s or after a total time of 2 minutes flushing (whichever occurs first) and document it.*
- ❒ Measure and record the amount of time required to reach steady temperature
- ❒ Measure and record water temperature at steady state
- ❒ Continuing flushing the hot water <u>(meaning: do not turn off the water between temperature and flow rate measurements)</u> and measure the water flow rate: record the amount of time it takes to fill the 1000 mL (repeat to have 3 measurements in total)
- ❒ Flush cold water over the sampling bottle to bring it below 50 °C (if applicable, depending on hot water temperature) to cool the sample down sufficiently such that it is not disinfected during transport.

<u>Second used shower (if applicable); collection of first liter from shower **through the showerhead**; cold and warm water mixed</u>

- ❒ If the shower head is too large to direct all the water into the 1-L wide-mouth glass bottle, use a UV irradiated plastic bag to act as a funnel to direct the water into the sampling bottle
- ❒ With the shower head (or funnel if needed) directed into an open 1-L wide-mouth glass bottle, open the cold and hot outlet(s) to get approximately equal proportion of cold and hot water flowing through the outlet and fill the bottle

  *Note: Open the outlet to medium flow rate (it should take approx. 5 s to fill the sample bottle)*
- ❒ Switch the shower head from the 1 L sample bottle to the 1000 mL beaker and collect 250 mL (you may quickly turn off the water in between)
- ❒ Turn off the fixture
- ❒ Firmly close the sampling bottle, turn bottle twice upside down to mix the thiosulfate
- ❒ Read and record sample temperature in the beaker with 250 mL (highest temperature that is reached on the display)

<u>Additional measurements at the kitchen tap; constant hot water temperature and flow rate</u>

- ❒ Turn on the hot water of the kitchen tap to the maximum flow rate. Continue flushing hot water until water temperature no longer changes.

  *Note: Flush the hot water until it is not changing by more than 0.1 °C for approx. 20 s or after a total time of 2 minutes flushing (whichever occurs first) and document it.*
- ❒ Measure and record the amount of time required to reach steady temperature
- ❒ Measure and record water temperature at steady state
- ❒ Continuing flushing the hot water <u>(meaning: do not turn off the water between temperature and flow rate measurements)</u> and measure the water flow rate: record the amount of time it takes to fill the 1000 mL (repeat to have 3 measurements in total)

<u>After sampling</u>

Samples will be processed within 24 hours.
